# Comparing neuromodulation targets to reduce cigarette craving and withdrawal: A randomized clinical trial

**DOI:** 10.1101/2024.10.10.24315204

**Authors:** Nicole Petersen, Michael R. Apostol, Timothy Jordan, Thuc Doan P. Ngo, Nicholas Kearley, Edythe D. London, Andrew Leuchter

## Abstract

**Importance:** Cigarette smoking remains the leading preventable cause of death worldwide, leading to development of new therapeutics, such as repetitive transcranial magnetic stimulation (rTMS).

**Objective:** We compared three TMS targets to evaluate the effects on cigarette craving and withdrawal, and on within- and between-network connectivity of the default mode, salience, and executive control networks.

**Design:** Data were collected using a repeated-measures, crossover trial. Investigators were not blinded, nor were participants, who were aware of the location of the stimulating magnet but not which locations were designated as control and experimental sites.

**Setting:** Data were collected in a neuromodulation clinic within an academic medical center.

**Participants:** Participants were men and women (44%) aged 21-45 (M = 33.3 years), who met DSM-5 criteria for tobacco use disorder and endorsed daily smoking for at least one year.

**Interventions:** TMS was delivered to the dorsolateral prefrontal cortex (dlPFC), superior frontal gyrus (SFG), and posterior parietal cortex (PPC). Area v5 of the visual cortex served as an active control site. Participants were scanned with resting-state fMRI and completed behavioral assessments before and after TMS.

**Main Outcomes and Measures:** Self-reports of craving, withdrawal, and negative affect were obtained, and resting-state functional connectivity of three canonical networks (executive control, default mode, and salience networks) was measured.

**Results:** Seventy-two participants completed at least one data collection session, and 57 completed all 4, yielding 61, 60, 62, and 66 complete stimulation sessions to the dlPFC, SFG, PPC, and v5, respectively. Stimulation to the SFG significantly reduced craving (95% CI, 0.0476-7.9559) and withdrawal (95% CI, 0.9225-8.1063) more than control stimulation. Effect sizes were larger in men (up D = 0.59) than in women (up to D =0.30). Neither PPC nor control site stimulation produced significant effects on craving, withdrawal, or negative affect. Functional connectivity analyses revealed that SFG stimulation did not produce significant changes to the networks examined, whereas dlPFC stimulation led to increased connectivity between somatomotor, default mode, and dorsal attention networks.

**Conclusions and Relevance:** The SFG appears to be a viable target for smoking-cessation treatment, especially for men, with possible advantages over dlPFC.

**Trial Registration:** Clinicaltrials.gov identifier: <u>NCT03827265</u>

**Question:** What is the most promising cortical target for TMS treatment of tobacco use disorder for men and women?

**Findings:** In a randomized crossover trial, stimulation to the superior frontal gyrus relieved craving and withdrawal the most.

**Meaning:** The superior frontal gyrus is a promising neuromodulation target for smoking cessation. Men and women may respond differently to this intervention.

## **I.** Introduction

Cigarette smoking is the leading preventable cause of death worldwide, despite substantial decreases in the prevalence of smoking, and novel therapeutics. The FDA recently cleared deep transcranial magnetic stimulation (TMS) for short-term smoking cessation, recognizing TMS as a safe and effective treatment for Tobacco Use Disorder^1^. However, evidence suggests that conventional figure-8 TMS coils may also be effective, and FDA approval of such a protocol would increase patient access by using TMS devices already available in clinics.

At least ten independent studies have demonstrated that figure-8 TMS reduces cigarette craving^2–11^, a predictor of substance use and relapse^12^. Conventional TMS has also led to reductions in heaviness of smoking,^2,3,8,11,13,14^ and improved smoking cessation outcomes^8,15–17^. Most studies have applied 10-20Hz stimulation to the left dorsolateral prefrontal cortex (dlPFC), but novel targets revealed by neuroimaging may optimize protocols. Increasingly, such targets appear to be multidimensional network targets rather than unitary locations^18^, and even adjacent unitary targets can be stimulated by TMS to influence unique networks^19^.

One model proposed that the salience network may act as an attentional “switch” during acute craving, allocating neural resources between the default mode and executive control networks^20,21^. Network coupling shifts after cigarette smoking, and the magnitude of these shifts corresponds to the magnitude of craving relief^22^.

Therefore, we sought to relieve craving by stimulating strategic nodes of these networks. We identified surface-level targets in each network accessible with conventional TMS, and hypothesized that stimulating each target would influence (1) craving and withdrawal symptoms, (2) connectivity within the targeted network, and, (3) connectivity between the targeted network and other canonical networks. Our primary objective was to identify the most effective protocol for reducing cigarette craving and withdrawal, and our secondary objective was to investigate the network perturbations produced by stimulating each target. We selected the dlPFC as an executive control network node, the superior frontal gyrus (SFG) as a salience network node, and the posterior parietal cortex (PPC) as a default mode network node.

Considering sex differences in nicotine withdrawal^23^ and neural correlates of craving,^24–26^ we also planned to determine whether the most effective stimulation target for men and women differed.

## **II.** Methods

The study protocol was approved by the UCLA Institutional Review Board UCLA (Medical IRB 3; #18-000509; initial approval: 07/27/2018). All participants provided informed consent in accordance with the Declaration of Helsinki. The clinical trial protocol is registered on clinicaltrials.gov, NCT03827265.

All analysis code is available at https://github.com/HumanBrainZappingatUCLA/TMS4TUD/.

### II.A. Study Design

Data were collected in a repeated measures, crossover design. Participants were asked to arrive on each testing day abstinent from smoking for >12 hours. Neuroimaging and behavioral measurements were collected before and after TMS to the three experimental sites (dlPFC, SFG, PPC) and the control site (area v5). The order of stimulation sites was randomized using a random number generator.

Investigators were not blinded at any stage. Participants could not be blinded to the location of the stimulating magnet, but were not informed which locations were control vs. experimental sites.

The trial’s planned stopping point was *N* = 60 based on an *a priori* power analysis (effect size estimate derived from unpublished data); 171 individuals were enrolled into the trial, and the total number of sessions for each target were:

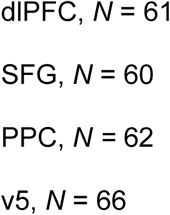

### II.B. Participants

Participant demographics are in Supplemental Table S1. Participants were recruited from the greater Los Angeles community via Craigslist and fliers. All data were collected in the Semel Institute for Neuroscience and Human Behavior at UCLA. A CONSORT diagram is shown in Figure 1.

**Figure 1:**
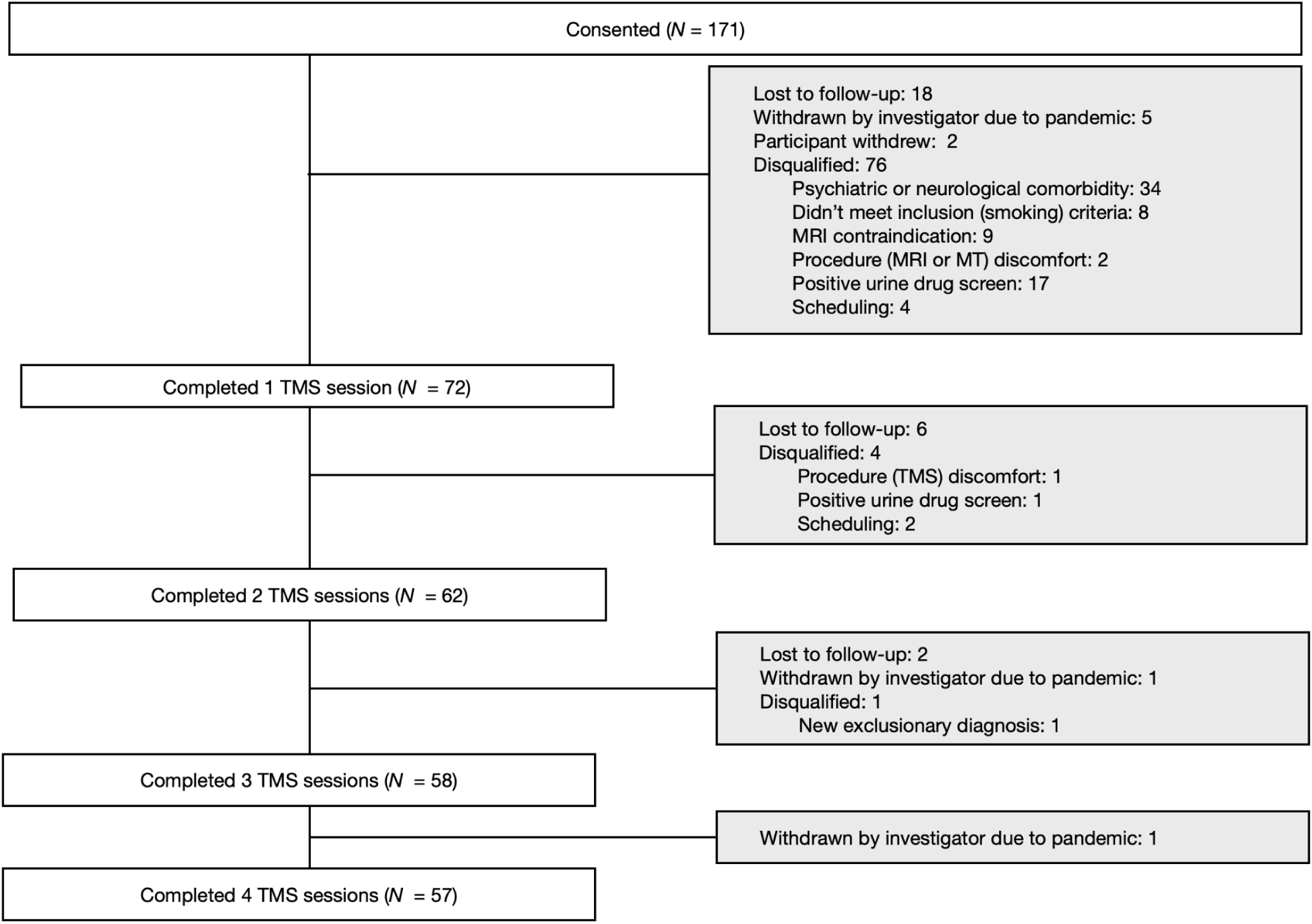
CONSORT Diagram. After initial telephone screening, 171 participants were consented for additional in-person screening. Slightly over half (55%) met eligibility criteria. Of these, 10% were lost to ordinary attrition, 3% were not scheduled due to COVID research shutdowns, and 1% withdrew after consenting, leading to 72 participants completing at least 1 TMS session. Of these, 80% would go on to complete all four TMS sessions.

Initial eligibility screening was conducted by phone, followed by an in-person session (if qualified). Smoking status was verified through expired carbon monoxide (Micro+ Smokerlyzer®breath CO monitor, Bedfont Scientific Ltd., Maidstone, Kent, UK) and urinary cotinine tests (Abbott™ NicQuick™ Nicotine/Cotinine Test or Accutest, Jant Pharmacal Corp., Encino, CA, USA), and abstinence from other substances was confirmed via urinalysis (Alere Toxicology Services, Portsmouth, VA, USA or Abbott™ iCup™ Zero Exposure Urine Drug Screen) and breathalyzer (Alco-Sensor FST breathalyzer; Intoximeters, Inc.). Comorbid psychiatric disorders were assessed using the Mini International Neuropsychiatric Interview^27^. Participants completed safety questionnaires to ensure eligibility for neuroimaging and neuromodulation. Inclusion criteria required meeting DSM-5 criteria for tobacco use disorder, >1 year of smoking history, daily use >4 cigarettes, and a positive urinary cotinine test. The age range was limited to 18-45 to control for potential age-related interactions with sex differences (e.g., menopause^28–30)^.

Participants were excluded if they had a recent (past six months) or current substance use disorder (except mild cannabis use), ongoing psychiatric conditions, major medical conditions, or were pregnant/breastfeeding. Exclusionary psychiatric diagnoses included major depressive disorder, bipolar I/II, anxiety disorders, PTSD, psychotic disorders, eating disorders, and antisocial personality disorder. Exclusion criteria also included medical conditions affecting major organ systems, positive tests for illicit substances (except THC), no biochemical verification of smoking (expired CO < 1 or negative cotinine), MRI/TMS safety risks (e.g., metal implants, seizures), active smoking cessation treatment, or left-hand dominance.

### II.C. Procedures

#### II.C.1. Behavioral measurements

Participants completed the Positive and Negative Affect Schedule (PANAS), Shiffman-Jarvik Withdrawal Questionnaire^31^ (SJWS; primary withdrawal measurement), and the Urge to Smoke scale^32^ (UTS; primary craving measurement); the latter assessments reliably capture symptoms of craving and withdrawal. All sessions took place in a quiet, controlled, private environment before and after TMS.

#### II.C.2. Brain imaging

Brain imaging data were collected on a 3-Tesla Siemens Prisma Fit MRI scanner with a 32-channel head coil before and immediately after TMS on each test day. A structural T1-weighted scan (TE=2.24 ms; TR=2400ms; isotropic voxels= 0.8mm^3^) and then functional T2*-weighted multi-band sequence (TE=37ms; TR=800ms; isotropic voxels=2mm^3^, volumes = 588) were collected.

FSL tools were used to apply motion correction, slice-timing correction, and normalization (FEAT, FMRI Expert Analysis Tool). ICA-FIX reduced noise and artifacts. Data were parcellated as in^33^, integrating the Schaefer 400-region cortical parcellation^34^ with 16 subcortical and 3 cerebellar regions, yielding 419 nodes.

Time series were extracted from these parcels, which were assigned to the default mode network, salience network, and executive control network via the Yeo 7-network solution^35,36^.

#### III.C.3. TMS

TMS was administered by a licensed physician at the UCLA Neuromodulation Division.

Two devices were used: A Magstim Super Rapid2 Plus1 system equipped with visor2 neuronavigation system (ANT Neuro), and a Magventure Magpro X100 with a Cool-B65 coil with Rogue Research Brainsight neuronavigation. Each stimulation dose was identical: 3,000 pulses of 10Hz stimulation were in 50 pulse trains of 5 seconds on, 10 seconds off (total = 15 minutes). Stimulation intensity was titrated to 100% of motor threshold (MT), with MT determined at the first treatment session as previously described^30,37,38^.

#### III.C.4. TMS Targeting

To personalize TMS targets based on resting-state functional connectivity, we developed a pipeline to identify the voxel within each target region (dlPFC, SFG, PPC, and v5) with maximum connectivity to key brain networks (executive control, salience, default mode, and visual). Preprocessed resting-state data were decomposed into 20 independent components using FSL’s MELODIC. Network hubs (posterior cingulate for default mode, inferior frontal gyrus for executive control, insula for salience, and area v5 for visual) were used to assign each component to a specific network. The TMS target was the voxel within each ROI with peak connectivity to the respective network.

### II.D. Outcome measures and statistical analysis

Self-reported cigarette craving, withdrawal, and negative affect during abstinence were measured using the Urge to Smoke (UTS) scale, Shiffman-Jarvik Withdrawal scale (SJWS), and the negative affect subscale of the Positive and Negative Affect Schedule (PANAS-), each administered before and after each TMS session. Linear mixed models (LMMs) were used to estimate the effect of TMS on each behavioral outcome using the smf.mixedlm function from the *statsmodels* library in Python, with the Restricted Maximum Likelihood (REML) method for model fitting. For each measure (UTS, SJWS, PANAS-), a separate LMM was fit to compare behavioral responses after stimulation at each experimental site (dlPFC, SFG, PPC) versus the control site (the v5 region of visual cortex). Each model included fixed effects for the time point (Pre/Post) and target group (Experimental/Control), as well as their interaction, and random intercepts for participants to account for within-subject correlations. The effect of sex on each primary outcome measurement was tested using a LMM, with participants entered as a random effect and sex entered as a fixed effect. All baseline (pre-TMS) data were included in the model.

### II.E. Brain imaging analysis

To analyze the effects of stimulating each neural target on functional connectivity, we calculated within- and between-network connectivity, initially focusing on the default mode network, salience network, and executive control network^35,36^. Connectivity values for network pairs were computed and averaged, then entered into difference scores. All statistics were carried out in Python (version 3.9.6) with the libraries *numpy*, *pandas*, *scipy*, and *statsmodels* implemented as needed. Paired *t*-tests compared pre- and post-session connectivity for each network pair, grouped by target. Next, LMMs were fitted in to model average connectivity as a function of time (pre/post, fixed effect) and target (fixed effect) while accounting for variability between participants (random effects). This was applied to the networks named above, for which we had *a priori* hypotheses, and also the remaining networks in the parcellation (limbic, dorsal attention, subcortical, visual, and somatomotor networks). Because all possible network pairs were included in the analysis, a familywise error correction was applied (false discovery rate using the Benjamini-Hochberg procedure).

## **III.** Results

### A.1. Establishing a stable baseline and adequacy of control

Baseline (pre-TMS) measurements of craving (UTS), withdrawal (SJWS), and negative affect (PANAS-) were stable across the four days of testing sessions (ps = 0.8477, 0.8887, and 0.7924, respectively), adding confidence that the washout period was adequate, and randomization successful.

Withdrawal (*β* = -0.039, SE = 1.193, *z* = -0.033, *p* = 0.974) and craving (*β* = 0.841, SE = 1.156, *z* =0.727, *p* = 0.467) did not change significantly or marginally from TMS delivered to v5 (control target). Negative affect also did not change significantly from TMS to this target (*β* = 1.246, SE = 0.649, *z* =1.919, *p* = 0.055), a strong trend was observed. *Post hoc* testing of individual PANAS- items showed that this effect was primarily driven by reduction in ratings of the “nervous” item (*p* = 0.0014) after stimulation.

Network connectivity was also similar at baseline between sessions. Within-network connectivity of limbic, dorsal attention, executive control, subcortical, visual, salience, somatomotor, and default mode networks was calculated and compared between test days. Comparing the connectivity of each baseline network-network pair by ANOVA did not yield any significant differences, *p*s ranging from 0.346534 (salience-salience connectivity) to 0.995908 (dorsal attention network-limbic network connectivity. Baseline network connectivity is shown in Supplemental Figure S2.

## **A.2.** Effect of sex on craving, withdrawal, affect, and network connectivity at baseline

Men and women did not differ significantly in craving (*p* = 0.4858), withdrawal (*p* = 0.1510), or negative affect (*p* = 0.3182). Baseline functional connectivity differed between men and women (Supplemental Table 1 and Supplemental Figure S3).

## **A.3.** Stimulation targets

The stimulation coordinates were clustered throughout each stimulation target. The distribution of these stimulation targets is shown in Supplemental Figure S2.

**B. Primary efficacy outcomes:** All results are visualized in raincloud plots (Figure 2).

**Figure 2:**
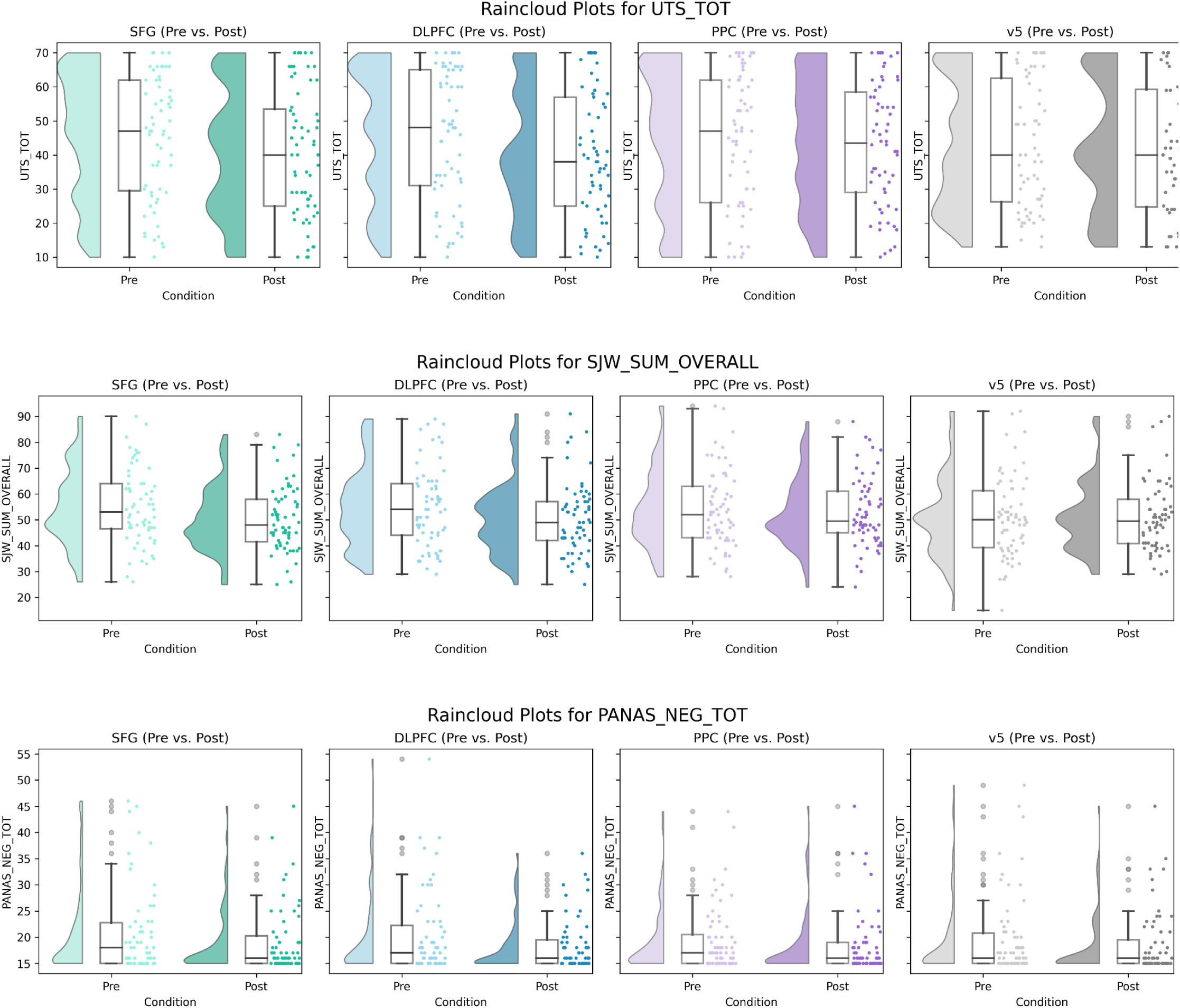
Raincloud plots depict the change in Urge to Smoke scale (UTS_TOT) scores (top panel), Shiffman-Jarvik Withdrawal scale scores (middle panel), and Negative Affect scores (bottom panel) from pre- to post-stimulation for the four stimulation targets. Each plot combines a density curve (left) indicating the distribution of scores, individual data points (middle) showing individual scores for pre- and post-stimulation conditions, and boxplots (right) summarizing the median and spread of the data.

**B.1. Craving:** A significant time-by-site interaction was found for stimulation to the SFG (β = 4.0018, SE = 2.0175, t = 1.9835, p = 0.0473) but not for dlPFC vs. control (β = 3.5579, SE = 2.2419, t = 1.5870, p = 0.1125) or PPC vs. control (β = 0.9769, SE = 2.2378, t = 0.4365, p = 0.6624) (Figure 2A).

**B.2. Withdrawal:** A significant time-by-site interaction was found for stimulation to the SFG (*β* = 4.5144, SE = 1.8326, *t* = 2.4633, *p* = 0.0138), but not for dlPFC vs. control (*β* = 3.4161, SE = 1.8632, *t* = 1.8335, *p* = 0.0667) or PPC vs. control (*β* = 1.4730, SE = 1.9197, *t* = 0.7673, *p* = 0.4429) (Figure 2B).

**B.3. Negative Affect:** No significant main effects on negative affect or time-by-target interactions on negative affect were found as a result of stimulation delivered to any target (Figure 2C) (dlPFC vs. control: *β* = 0.5643, SE = 0.9950, *t* = 0.5671, *p* = 0.5706; SFG vs. control*: β* = 0.6511, SE = 1.1722, *t* = 0.5555, *p* = 0.5786; PPC vs. control*: β* = -0.2524, SE = 0.9485, *t* = -0.2661, *p* = 0.7902).

## **C.** Potential confounds

Fagerström Test for Nicotine Dependence (FTND) scores were related to craving (*p* = 0.00033035), withdrawal (*p* = 0.000031969), and negative affect (*p* = 0.018471), but did not interact significantly with the effect of time on any variable (all interaction *p*s > 0.24). Age was unrelated to dependent variables variables (*p*s > 0.10).

Years of education was related to baseline psychological withdrawal, *p*_uncorrected_ = 0.04. Therefore, these variables were excluded from subsequent models.

## **D.** Sex differences

Three-way interactions between sex, target, and time on craving, withdrawal, and negative affect were not significant (*p*s > 0.05). However, the time-by-sex interaction on overall withdrawal was significant for SFG stimulation only (*p* = 0.0395).

## E. *Post hoc* data exploration

Change scores showing the effect of each stimulation type on unused subscales of primary analysis instruments (Shiffman-Jarvik Withdrawal Scale: Craving, Psychological Withdrawal, Physiological Withdrawal, Stimulation/Sedation, and Appetite; PANAS: positive affect) are shown in Figure 3. The largest effects were craving reductions from SFG and dlPFC stimulation, followed by reductions in withdrawal as a result of SFG stimulation.

**Figure 3:**
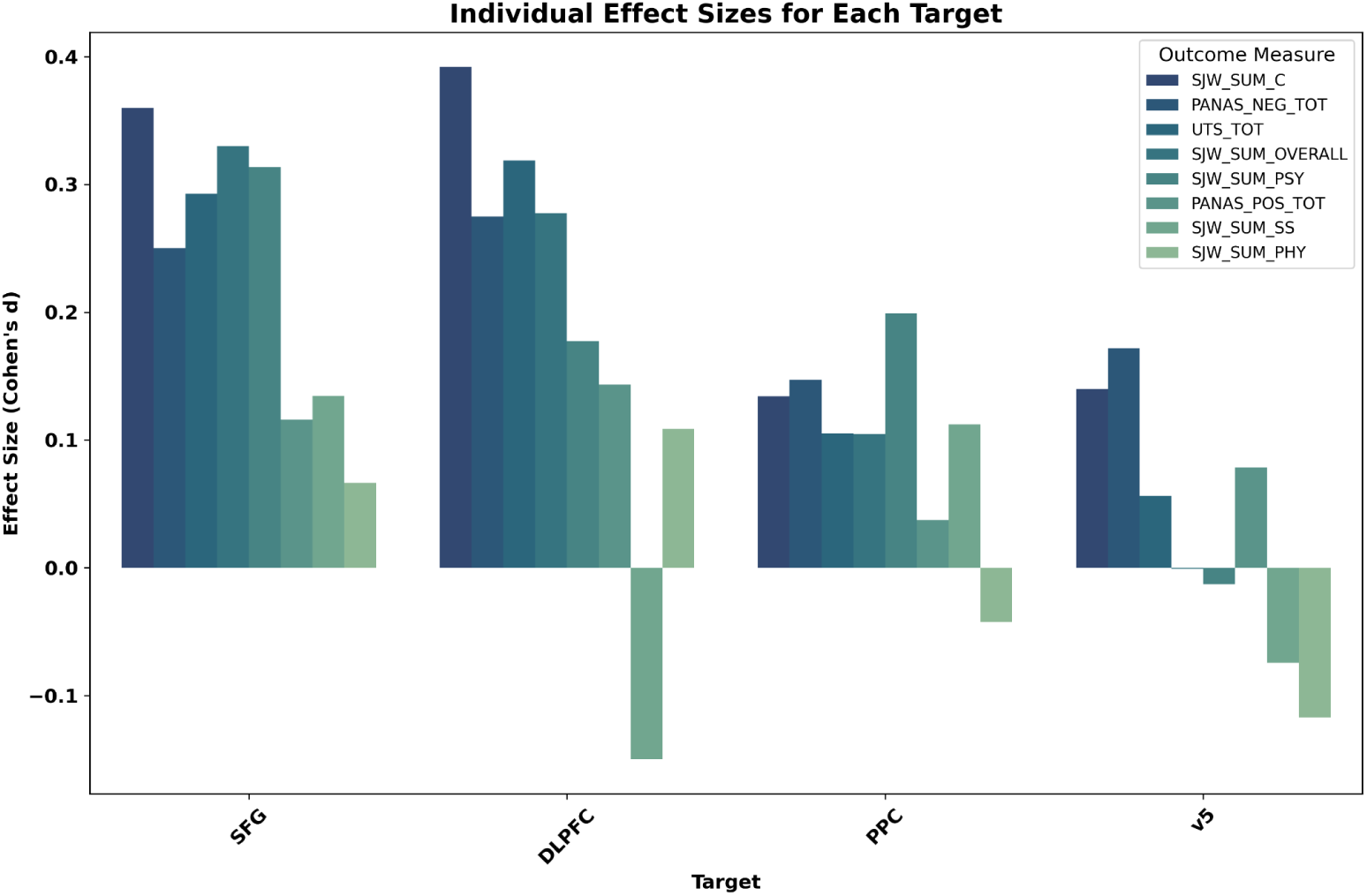
Effect sizes showing TMS’ effect on each inventory’s subscales, separated by stimulation target. Stimulation to SFG and dlPFC produced some medium-sized effects, whereas stimulation to PPC and v5 led to no or small effects. SJW_SUM_C = Shiffman-Jarvik craving subscale; PANAS_NEG_TOT = Positive and Negative Affect Schedule, negative affect subscale; UTS_TOT = Urge to Smoke Total; SJW_SUM_OVERALL = Shiffman-Jarvik Withdrawal Scale total score; SJW_SUM_PSY = Shiffman-Jarvik Withdrawal scale, psychological withdrawal subscale; PANAS_POS_TOT = Positive and Negative Affect Schedule, positive affect subscale; SJW_SUM_SS = Shiffman-Jarvik Withdrawal Scale stimulation/sedation subscale; SJW_SUM_PHY = Shiffman-Jarvik Withdrawal Scale, physiological withdrawal subscale

Disaggregating by sex, SFG stimulation had larger effects in men than women (Figure 4). The largest effect in the dataset for men was approximately twice as large as the largest effect size for women, suggesting that men drive the effects of SFG stimulation on craving and withdrawal in the overall model.

**Figure 4:**
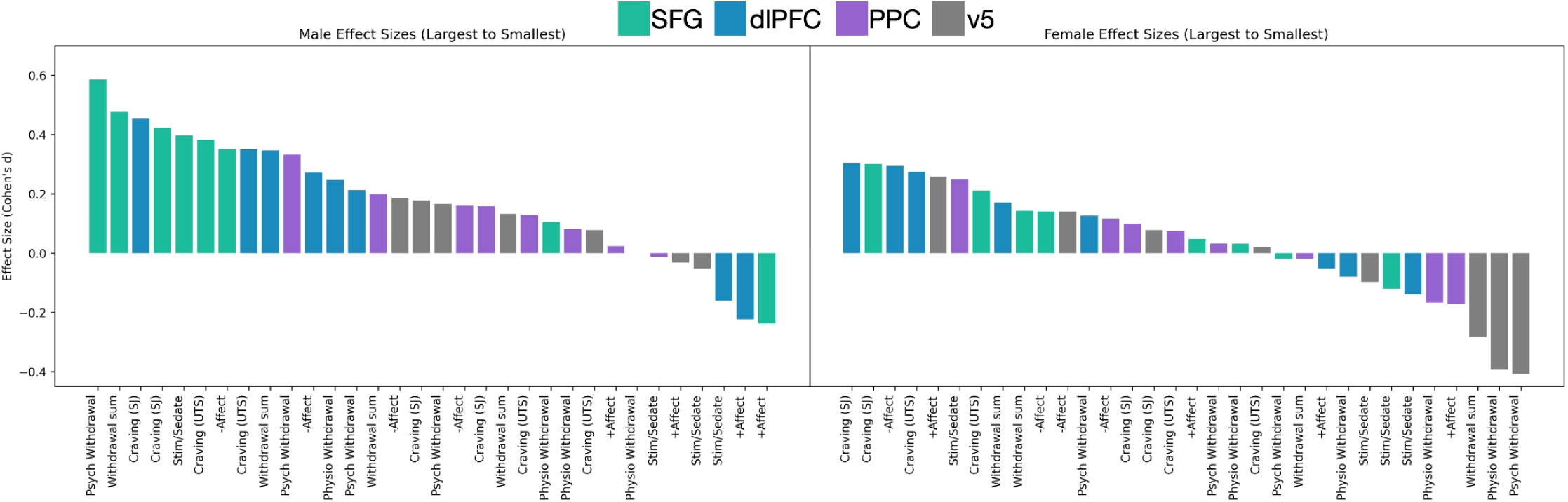
Effect sizes of each stimulation type separated by sex. The pattern of effect sizes appears different for men and women. Men show a small-medium change (reduction) in craving and withdrawal as a response to TMS to the SFG and dlPFC. The largest effect in the trial is a reduction in psychological withdrawal resulting from SFG stimulation in men only. Women show smaller effects overall, especially with respect to reductions in psychological withdrawal.

Exploratory tests for *main* effects of stimulation indicated that TMS to dlPFC reduced craving (11.64% decrease, *p* = 0.0053), as did SFG TMS (10.69% decrease, *p* = 0.0013), but not v5 (1.97% decreas, *p* = 0.47) nor PPC (4.10% reduction, p = 0.186).

## G. Safety outcomes

Safety outcomes are detailed in Supplemental Table S2. Two participants withdrew from the study due to discomfort from the stimulation procedures, both on days randomized to dlPFC stimulation. There were no study-related severe or unexpected adverse events.

## H. Brain imaging findings

Planned imaging analyses focused on the default mode, executive, and salience networks (Figure 5). Connectivity within and between these networks was not significantly different after TMS to any target, nor did the time-by-target interaction, all *p*s > 0.05.

**Figure 5:**
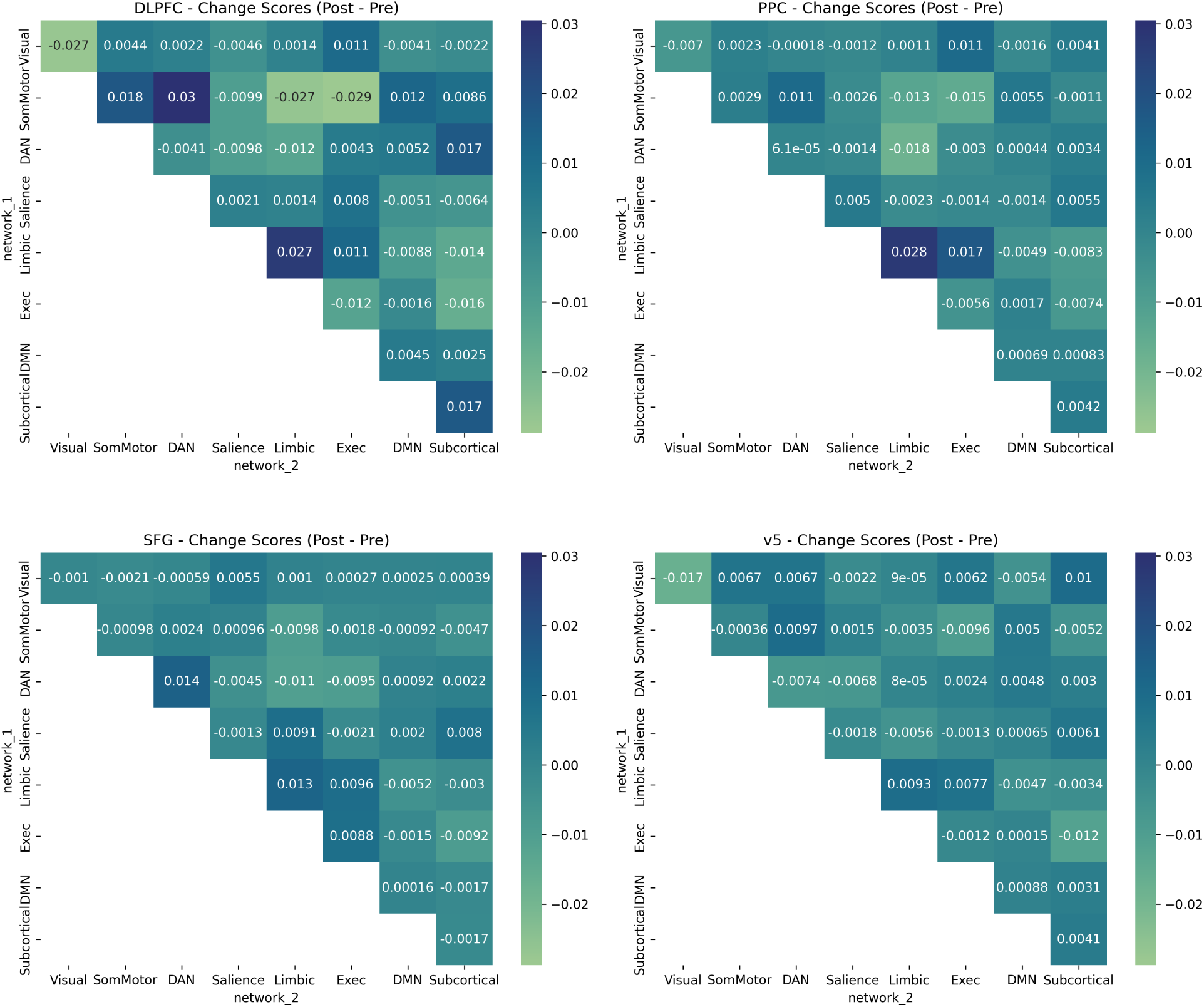
Effects of TMS on canonical network connectivity. Only TMS to dlPFC produced changes in network connectivity that survived familywise error correction. These changes included both increases in connectivity (between the somatomotor network and the default mode network, and between the somatomotor network and dorsal attention network), and decreases in connectivity (within the visual network, and between the somatomotor network and executive control network).

Exploratory analysis of all networks revealed that dlPFC stimulation only changed network connectivity.

After dlPFC TMS, connectivity was significantly **higher**:

1. Between the somatomotor network and default mode network (*p*_FDR_ = 0.030)
2. Between the somatomotor network and dorsal attention network (*p*_FDR_ = 0.022) After dlPFC TMS, connectivity was significantly **lower**:
3. Within the visual network (*p*_FDR_ = 0.022)
4. Between the somatomotor network and executive network (*p*_FDR_ = 0.011)

## Discussion

In this randomized crossover trial evaluating the acute effects of single-session TMS delivered to three experimental sites (dlPFC, SFG, PPC) and a control site, stimulation to the SFG significantly reduced both craving and withdrawal in individuals with Tobacco Use Disorder compared to the control target, replicating earlier findings^9^. Negative affect was not significantly impacted by stimulation to any target as compared to the control target. Our findings are also consistent with previous evidence that stimulation to the dlPFC reduces craving^2–4,6,8,10,11^. The effect sizes for men were larger than the effect sizes for women. SFG is a promising target for smoking cessation trials and can be implemented using standard TMS equipment.

We selected stimulation targets that are hubs of resting-state networks (dlPFC for executive control, SFG for salience, PPC for default mode, and v5 for visual), expecting network-specific effects. However, dlPFC was globally influential, while SFG and v5 produced no significant network-level changes. Nonetheless, SFG stimulation effectively reduced craving and withdrawal, suggesting alternative neural mechanisms. Future research could search for circuits that underlie the craving reductions observed, and incorporate them into network-guided studies, similar to success in network-guided TMS for depression treatment^39–43^.

Symptom-specific targeting is also emerging for depression treatment^44^, and our finding that craving and withdrawal may respond to different targets suggests that this is a potential avenue for addiction treatment as well.

This trial was impacted by the COVID-19 pandemic, with data collection spanning from March 2019 to August 2024. During this time, changes such as increased use of electronic nicotine delivery systems and cannabis may have influenced results. Additionally, our exclusion of participants with comorbid psychiatric and substance use disorders limits generalizability, as TMS may be effective in treating such patients^45^. The trial was also underpowered to detect sex differences, likely due to the higher demands of detecting ordinal interactions^46^. Finally, the relatively low dose of neuromodulation may have limited the separation between experimental and control conditions, as higher doses are typically used in treatment protocols.

Phase 2 and 3 trials are needed to confirm the efficacy of SFG stimulation for smoking cessation, followed by optimization trials examining factors such as stimulation frequency, dosing, spacing, and neural context. Additionally, future studies should compare the tolerability of different TMS protocols. Tailoring addiction treatment based on individual symptom profiles could also be explored, with different neural targets emerging for individuals whose relapse risk is driven by craving, withdrawal, or negative affect. Investigating symptom-specific neural targets may help overcome observed sex differences and other individual-level factors that influence brain circuits related to craving and withdrawal relief.

## Data Availability

All data produced in the present study are available upon reasonable request to the authors.

## Acknowledgments, funding, and disclosures

A number of people contributed to this manuscript. Andrea Donis, Kaitlin Kinney, Maylen Perez Diaz, Graham Pilger, Anthony Sun, Dora Ambrose, Gino Haase, Ishaa Diwakar, Jianna Ursitti, Lauren Kim, Malia Belnap, Melanie Beltran, Riley Russell, Brian Bossé, and Cole Matthews assisted in data collection. Jed Rose and Steve Shoptaw contributed to study conceptualization and design.

## Funding

This work was supported by K99/R00DA045749 (*Transcranial Magnetic Stimulation and Tobacco Use Disorder: A Network-Level Approach with Attention to Sex as a Biological Variable*) and the Friends of Semel Award to NP.

Artificial intelligence technologies: OpenAI GPT-4.0 was used to assist in writing and debugging analysis code.

## References

1. Zangen A, Moshe H, Martinez D, et al. Repetitive transcranial magnetic stimulation for smoking cessation: a pivotal multicenter double-blind randomized controlled trial. World Psychiatry. 2021;20(3):397–404. doi:10.1002/wps.20905

2. Abdelrahman AA, Noaman M, Fawzy M, Moheb A, Karim AA, Khedr EM. A double-blind randomized clinical trial of high frequency rTMS over the DLPFC on nicotine dependence, anxiety and depression. Scientific Reports. 2021;11(1):1640. doi:10.1038/s41598-020-80927-5

3. Amiaz R, Levy D, Vainiger D, Grunhaus L, Zangen A. Repeated high-frequency transcranial magnetic stimulation over the dorsolateral prefrontal cortex reduces cigarette craving and consumption. Addiction. 2009;104(4):653–660.

4. Chang D, Zhang J, Peng W, et al. Smoking cessation with 20 Hz repetitive transcranial magnetic stimulation (rTMS) applied to two brain regions: a pilot study. Frontiers in human neuroscience. 2018;12:344.

5. Flores-Leal M, Sacristán-Rock E, Jiménez-Angeles L, Azpiroz-Leehan J. Low frequency repetitive transcranial magnetic stimulation effects over dorsolateral prefrontal cortex in moderate nicotine dependent subjects. In: VI Latin American Congress on Biomedical Engineering CLAIB 2014, Paraná, Argentina 29, 30 & 31 October 2014. Springer; 2015:317-320.

6. Johann M, Wiegand R, Kharraz A, et al. Transcranial magnetic stimulation for nicotine dependence. Psychiatrische Praxis. 2003;30:S129–31.

7. Li X, Hartwell KJ, Owens M, et al. Repetitive Transcranial Magnetic Stimulation of the Dorsolateral Prefrontal Cortex Reduces Nicotine Cue Craving. Biological Psychiatry. 2013;73(8):714–720. doi:10.1016/j.biopsych.2013.01.003

8. Li X, Hartwell KJ, Henderson S, Badran BW, Brady KT, George MS. Two weeks of image-guided left dorsolateral prefrontal cortex repetitive transcranial magnetic stimulation improves smoking cessation: A double-blind, sham-controlled, randomized clinical trial. Brain Stimulation. 2020;13(5):1271–1279. doi:10.1016/j.brs.2020.06.007

9. Rose JE, McClernon FJ, Froeliger B, Behm FM, Preud’homme X, Krystal AD. Repetitive Transcranial Magnetic Stimulation of the Superior Frontal Gyrus Modulates Craving for Cigarettes. Biological Psychiatry. 2011;70(8):794–799. doi:10.1016/j.biopsych.2011.05.031

10. Shevorykin A, Carl E, Mahoney MC, et al. Transcranial Magnetic Stimulation for Long-Term Smoking Cessation: Preliminary Examination of Delay Discounting as a Therapeutic Target and the Effects of Intensity and Duration. Frontiers in Human Neuroscience. 2022;16. https://www.frontiersin.org/journals/human-neuroscience/articles/10.3389/fnhum.2022.920383

11. Upton S, Brown AA, Ithman M, et al. Effects of Hyperdirect Pathway Theta Burst Transcranial Magnetic Stimulation on Inhibitory Control, Craving, and Smoking in Adults With Nicotine Dependence: A Double-Blind, Randomized Crossover Trial. Biological Psychiatry: Cognitive Neuroscience and Neuroimaging. 2023;8(11):1156–1165. doi:10.1016/j.bpsc.2023.07.014

12. Vafaie N, Kober H. Association of Drug Cues and Craving With Drug Use and Relapse: A Systematic Review and Meta-analysis. JAMA Psychiatry. 2022;79(7):641–650. doi:10.1001/jamapsychiatry.2022.1240

13. Eichhammer P, Johann M, Kharraz A, et al. High-frequency repetitive transcranial magnetic stimulation decreases cigarette smoking. Journal of Clinical Psychiatry. 2003;64(8):951–953.

14. Prikryl R, Ustohal L, Kucerova HP, et al. Repetitive transcranial magnetic stimulation reduces cigarette consumption in schizophrenia patients. Progress in Neuro-Psychopharmacology and Biological Psychiatry. 2014;49:30–35.

15. Dieler AC, Dresler T, Joachim K, Deckert J, Herrmann MJ, Fallgatter AJ. Can intermittent theta burst stimulation as add-on to psychotherapy improve nicotine abstinence? Results from a pilot study. European addiction research. 2014;20(5):248–253.

16. Sheffer CE, Bickel WK, Brandon TH, et al. Preventing relapse to smoking with transcranial magnetic stimulation: Feasibility and potential efficacy. Drug and Alcohol Dependence. 2018;182:8–18. doi:10.1016/j.drugalcdep.2017.09.037

17. Trojak B, Meille V, Achab S, et al. Transcranial magnetic stimulation combined with nicotine replacement therapy for smoking cessation: a randomized controlled trial. Brain stimulation. 2015;8(6):1168–1174.

18. Noble S, Curtiss J, Pessoa L, Scheinost D. The tip of the iceberg: A call to embrace anti-localizationism in human neuroscience research. Imaging Neuroscience. 2024;2:1–10. doi:10.1162/imag_a_00138

19. Eldaief MC, McMains S, Izquierdo-Garcia D, Daneshzand M, Nummenmaa A, Braga RM. Network-specific metabolic and haemodynamic effects elicited by non-invasive brain stimulation. Nature Mental Health. Published online 2023:1–15.

20. Fedota JR, Stein EA. Resting-state functional connectivity and nicotine addiction: prospects for biomarker development. Annals of the New York Academy of Sciences. 2015;1349(1):64–82. 10.1111/nyas.12882

21. Sutherland MT, McHugh MJ, Pariyadath V, Stein EA. Resting state functional connectivity in addiction: Lessons learned and a road ahead. Neuroimage. 2012;62(4):2281–2295.

22. Lerman C, Gu H, Loughead J, Ruparel K, Yang Y, Stein EA. Large-scale brain network coupling predicts acute nicotine abstinence effects on craving and cognitive function. JAMA Psychiatry. 2014;71(5):523–530. doi:10.1001/jamapsychiatry.2013.4091

23. Faulkner P, Petersen N, Ghahremani DG, et al. Sex differences in tobacco withdrawal and responses to smoking reduced-nicotine cigarettes in young smokers. Psychopharmacology. 2018;235(1):193–202. doi:10.1007/s00213-017-4755-x

24. Dumais KM, Franklin TR, Jagannathan K, et al. Multi-site exploration of sex differences in brain reactivity to smoking cues: Consensus across sites and methodologies. Drug and Alcohol Dependence. 2017;178:469–476. doi:10.1016/j.drugalcdep.2017.05.044

25. Perez Diaz M, Pochon JB, Ghahremani DG, et al. Sex Differences in the Association of Cigarette Craving With Insula Structure. International Journal of Neuropsychopharmacology. 2021;24(8):624–633. doi:10.1093/ijnp/pyab015

26. Wetherill RR, Young KA, Jagannathan K, et al. The impact of sex on brain responses to smoking cues: a perfusion fMRI study. Biology of Sex Differences. 2013;4(1):9. doi:10.1186/2042-6410-4-9

27. Sheehan DV, Lecrubier Y, Sheehan KH, et al. The Mini-International Neuropsychiatric Interview (M.I.N.I.): the development and validation of a structured diagnostic psychiatric interview for DSM-IV and ICD-10. J Clin Psychiatry. 1998;59 Suppl 20:22-33;quiz 34-57.

28. Kryatova MS, Seiner SJ, Brown JC, Siddiqi SH. Older age associated with better antidepressant response to H1-coil transcranial magnetic stimulation in female patients. Journal of Affective Disorders. 2024;351:66–73.

29. Sackeim HA, Aaronson ST, Carpenter LL, et al. Clinical outcomes in a large registry of patients with major depressive disorder treated with Transcranial Magnetic Stimulation. Journal of Affective Disorders. 2020;277:65–74. doi:10.1016/j.jad.2020.08.005

30. Slan AR, Citrenbaum C, Corlier J, et al. The role of sex and age in the differential efficacy of 10 Hz and intermittent theta-burst (iTBS) repetitive transcranial magnetic stimulation (rTMS) treatment of major depressive disorder (MDD). Journal of Affective Disorders. 2024;366:106–112. doi:10.1016/j.jad.2024.08.129

31. Shiffman SM, Jarvik ME. Smoking withdrawal symptoms in two weeks of abstinence. Psychopharmacology. 1976;50:35–39.

32. Jarvik ME, Madsen DC, Olmstead RE, Iwamoto-Schaap PN, Elins JL, Benowitz NL. Nicotine blood levels and subjective craving for cigarettes. Pharmacology Biochemistry and Behavior. 2000;66(3):553–558.

33. Van De Ville D, Farouj Y, Preti MG, Liégeois R, Amico E. When makes you unique: Temporality of the human brain fingerprint. Science advances. 2021;7(42):eabj0751.

34. Schaefer A, Kong R, Gordon EM, et al. Local-global parcellation of the human cerebral cortex from intrinsic functional connectivity MRI. Cerebral cortex. 2018;28(9):3095–3114.

35. Yeo BT, Krienen FM, Sepulcre J, et al. The organization of the human cerebral cortex estimated by intrinsic functional connectivity. Journal of neurophysiology. Published online 2011.

36. Yeo BTT, Krienen FM, Chee MWL, Buckner RL. Estimates of segregation and overlap of functional connectivity networks in the human cerebral cortex. NeuroImage. 2014;88:212–227. doi:10.1016/j.neuroimage.2013.10.046

37. Chu SA, Tadayonnejad R, Corlier J, Wilson AC, Citrenbaum C, Leuchter AF. Rumination symptoms in treatment-resistant major depressive disorder, and outcomes of repetitive Transcranial Magnetic Stimulation (rTMS) treatment. Translational Psychiatry. 2023;13(1):293. doi:10.1038/s41398-023-02566-4

38. Citrenbaum C, Corlier J, Ngo D, et al. Pretreatment pupillary reactivity is associated with differential early response to 10 Hz and intermittent theta-burst repetitive transcranial magnetic stimulation (rTMS) treatment of major depressive disorder (MDD). Brain Stimulation. 2023;16(6):1566–1571.

39. Cash RFH, Cocchi L, Lv J, Fitzgerald PB, Zalesky A. Functional Magnetic Resonance Imaging–Guided Personalization of Transcranial Magnetic Stimulation Treatment for Depression. JAMA Psychiatry. 2021;78(3):337–339. doi:10.1001/jamapsychiatry.2020.3794

40. Fox MD, Buckner RL, White MP, Greicius MD, Pascual-Leone A. Efficacy of Transcranial Magnetic Stimulation Targets for Depression Is Related to Intrinsic Functional Connectivity with the Subgenual Cingulate. Biological Psychiatry. 2012;72(7):595–603. doi:10.1016/j.biopsych.2012.04.028

41. Kong G, Wei L, Wang J, Zhu C, Tang Y. The therapeutic potential of personalized connectivity-guided transcranial magnetic stimulation target over group-average target for depression. *Brain Stimulation: Basic*, Translational, and Clinical Research in Neuromodulation. 2022;15(5):1063–1064. doi:10.1016/j.brs.2022.07.054

42. Oathes DJ, Gonzalez AF, Grier J, Blaine C, Garcia SD, Linn KA. Clinical Response to fMRI-guided Compared to Non-Image Guided rTMS in Depression and PTSD: A Randomized Trial. medRxiv. Published online January 1, 2024:2024.07.29.24311191. doi:10.1101/2024.07.29.24311191

43. Siddiqi SH, Weigand A, Pascual-Leone A, Fox MD. Identification of personalized transcranial magnetic stimulation targets based on subgenual cingulate connectivity: an independent replication. Biological Psychiatry. 2021;90(10):e55–e56.

44. Siddiqi SH, Taylor SF, Cooke D, Pascual-Leone A, George MS, Fox MD. Distinct Symptom-Specific Treatment Targets for Circuit-Based Neuromodulation. AJP. 2020;177(5):435–446. doi:10.1176/appi.ajp.2019.19090915

45. Blyth SH, Zabik NL, Krosche A, Prisciandaro JJ, Ward HB. rTMS for Co-occurring Psychiatric and Substance Use Disorders: Narrative Review and Future Directions. Current Addiction Reports. 2024;11(2):342–351. doi:10.1007/s40429-024-00542-6

46. Strube MJ, Bobko P. Testing hypotheses about ordinal interactions: Simulations and further comments. Journal of Applied Psychology. 1989;74(2):247.

