## Supplement for "Comparing neuromodulation targets to reduce cigarette craving and withdrawal: A randomized clinical trial"

**Supplemental Information:**


Page 1: Supplemental Table S1, Demographics

Page 2: Supplemental Table S2, Safety information

Page 3: Supplemental Figure S1, Baseline resting-state functional connectivity is consistent between sessions
Supplemental Figure S2, Distribution of stimulation sites

Page 4: Temporal spacing of sessions
Supplemental Table S3, Baseline sex differences in inter- and intra-network connectivity

Page 5: Supplemental Figure S3: Baseline sex differences and similarities in the connectivity of each network-network pair.

**Supplemental Table S1:** Demographic characteristics of 72 included study participants.

| | |
|---|---|
| **Age** | |
| *M* | 33.3 years |
| *SD* | 6.46 years |
| Range | 21 to 45 years |
| **Sex** | |
| Female | 32 (44.4%) |
| Male | 40 (55.6%) |
| **Gender** | |
| Female | 31 (43.0%) |
| Male | 40 (55.6%) |
| Prefer not to answer | 1 (1.4%) |
| **Race and ethnicity** | |
| White, non-hispanic | 28 (38.89%) |
| White, hispanic | 7 (9.72%) |
| Black or African American | 17 (23.61%) |
| Asian | 8 (11.11%) |
| More than One Race | 7 (9.72%) |
| Native Hawaiian or Other Pacific Islander | 2 (2.78%) |
| Unknown or Prefer Not to Specify | 3 (4.17%) |
| **Years of education** | |
| *M* | 14.2 years |
| *SD* | 1.95 |
| Range | 12 to 20 years |

*Safety information:*
Unrelated events: During the trial, one serious adverse event occurred. A participant informed the study team of a new cancer diagnosis, which was determined to be unrelated to the trial intervention (the cancer had no central nervous system involvement). The participant was under the care of their healthcare provider, and the study team subsequently withdrew them from the trial.

Separately, a participant with a history of migraine experienced a migraine 48 hours after the initial TMS session. This was deemed unlikely to be study related.

Mild, expected AEs that were or probably were study related are given in Table 2:

**Supplemental Table S2:** Mild, expected adverse events following stimulation to each site. The most frequent event was twitching, pain, or discomfort in the head or face.

| Event | Motor threshold measurement | dlPFC | SFG | PPC | v5 |
|---|---|---|---|---|---|
| **Twitching, pain, or discomfort in head or face** | 0 | 15 | 4 | 0 | 1 |
| **Twitching, discomfort, or pain in neck or shoulders** | 0 | 2 | 0 | 0 | 0 |
| **Twitching, discomfort, or pain in arms or hands** | 0 | 1 | 0 | 0 | 0 |
| **Drowsiness** | 0 | 1 | 2 | 1 | 2 |
| **Disorientation** | 0 | 2 | 1 | 0 | 1 |
| **Anxiety** | 0 | 0 | 1 | 0 | 0 |
| **Vasovagal reaction** | 1 | 0 | 0 | 0 | 0 |

**Supplemental Figure S1: Resting-state networks before TMS on each study day.** The average functional connectivity of the limbic, dorsal attention (DAN), executive control (Exec), subcortical, visual, salience, somatomotor (SomMotor), and default mode (DMN) networks are shown at baseline, before the delivery of TMS on each testing day. The order of stimulation to each site was randomized, and network connectivity is averaged by stimulation site. Average network connectivity of each network was similar at baseline.

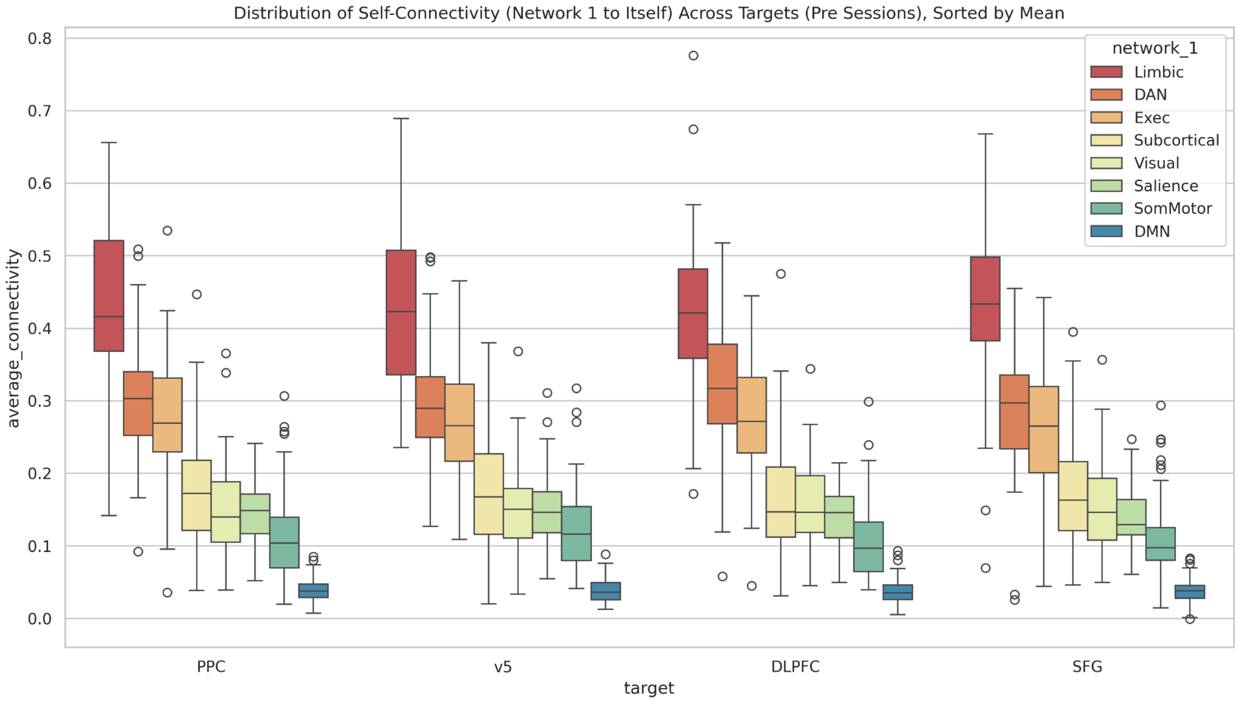


**Supplemental Figure S2: Spatial distribution of stimulation targets.** The specific voxel targeted within each ROI is shown below. Blue dots show the distribution of targets within the dlPFC, green show the distribution within the SFG, purple within the PPC, and gray within v5.

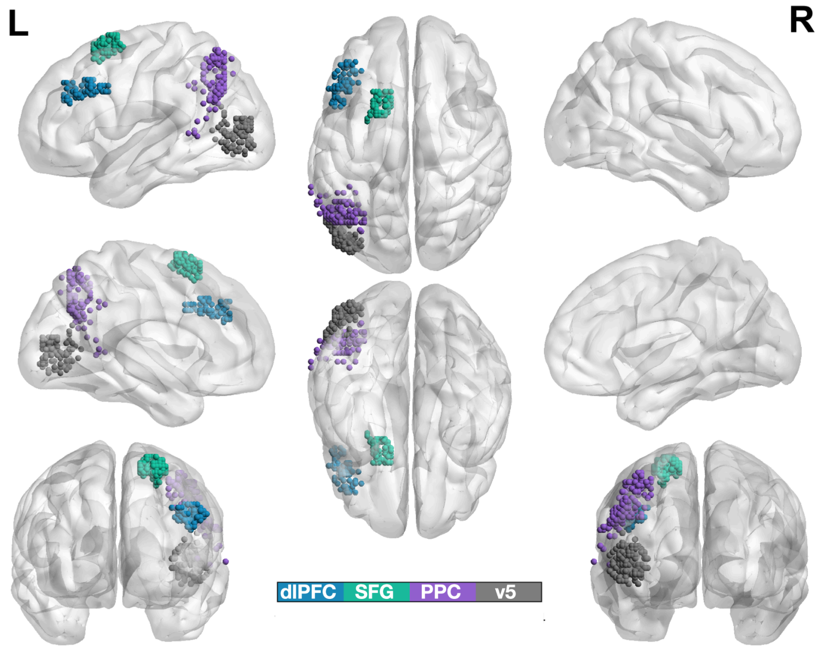

*Spacing between sessions*

The minimum amount of time between testing sessions was 24 hours. We did not impose a maximum; if participants were able to return for additional testing after a delay (illness, travel, other scheduling reasons), they were still included. Therefore, the number of days that elapsed between testing sessions ranged from 1 to 75, with a mean of 8.55 days. The median length of time between sessions was 5.01 days.

*Sex-specific effects on network connectivity*

Exploratory analyses showed a relatively small effect of sex in network responses to TMS, and a larger effect of sex on network connectivity at baseline. Disaggregating the data by sex, the only network-network connectivity change that survived FDR correction was the change in dorsal attention-subcortical network connectivity after dlPFC stimulation ($p_{FDR} = 0.043761$); *i.e.*, fewer effects of TMS were found when men and women were separated. No other reported network change survived FDR correction when men and women's data were tested separately.

We did observe notable differences in **baseline** network connectivity between men and women. Grouping all baseline data together, sex differences were observed in connectivity of the subcortical network to every other network except the visual network (executive, limbic, dorsal attention, somatomotor, default mode, and salience networks) at $p_{FDR} < 0.0005$). Higher connectivity in women (vs. men) was observed within the subcortical network, and between the subcortical and executive, limbic, and salience networks. Lower connectivity in women (vs. men) was observed between the subcortical network and the dorsal attention, somatomotor, and default mode networks.

Also surviving familywise error correction were sex differences in connectivity of the visual network to itself, the default mode network, and the dorsal attention network, as well as dorsal attention-dorsal attention network connectivity. These results are presented again below in supplemental Table S1. The average connectivity at baseline for every network-network pair is shown next (Supplemental Figure S1), disaggregated to show data separately for men and women.

**Supplemental Table S3: Baseline sex differences in inter- and intra-network connectivity.** *Abbreviations:* Exec = executive control network, DAN = dorsal attention network, SomMotor = somatomotor network, DMN = default mode network. Negative coefficients reflect lower connectivity in women.

| Network Pair | p-value | FDR-corrected p-value | Coefficient |
|---|---|---|---|
| Exec-Subcortical | 3.68e-08 | 0.000001 | -0.069482 |
| Limbic-Subcortical | 1.02e-06 | 0.000018 | -0.072067 |
| DAN-Subcortical | 5.31e-06 | 0.000064 | 0.052976 |
| SomMotor-Subcortical | 7.47e-06 | 0.000067 | 0.037885 |
| DMN-Subcortical | 1.16e-05 | 0.000083 | 0.022817 |
| Salience-Subcortical | 7.21e-05 | 0.000371 | -0.033027 |
| Subcortical-Subcortical | 6.69e-05 | 0.000371 | -0.057213 |
| Visual-Visual | 1.36e-04 | 0.000611 | -0.039785 |
| Visual-DMN | 2.95e-03 | 0.011786 | -0.013223 |
| DAN-DAN | 3.43e-03 | 0.012346 | -0.041229 |

| | | | |
|---|---|---|---|
| Visual-DAN | 9.51e-03 | 0.031133 | 0.020820 |

**Supplemental Figure S3: Baseline sex differences and similarities in the connectivity of each network-network pair**. The plots are arranged in no particular order. The y-axis is average connectivity. Data are disaggregated by sex, with women shown in purple and men in blue. The general pattern demonstrates that the observed sex differences and similarities are consistent between testing days.

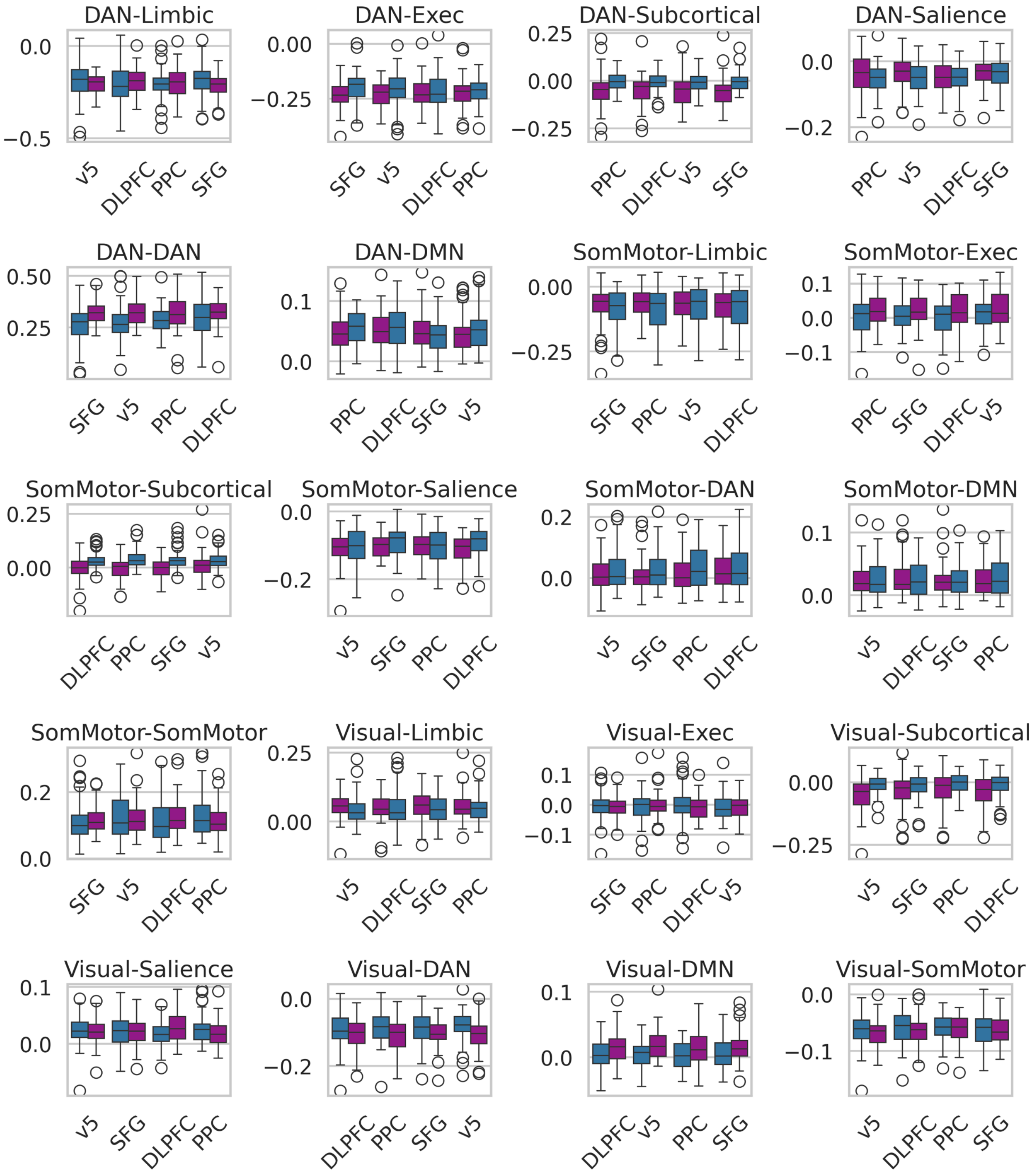

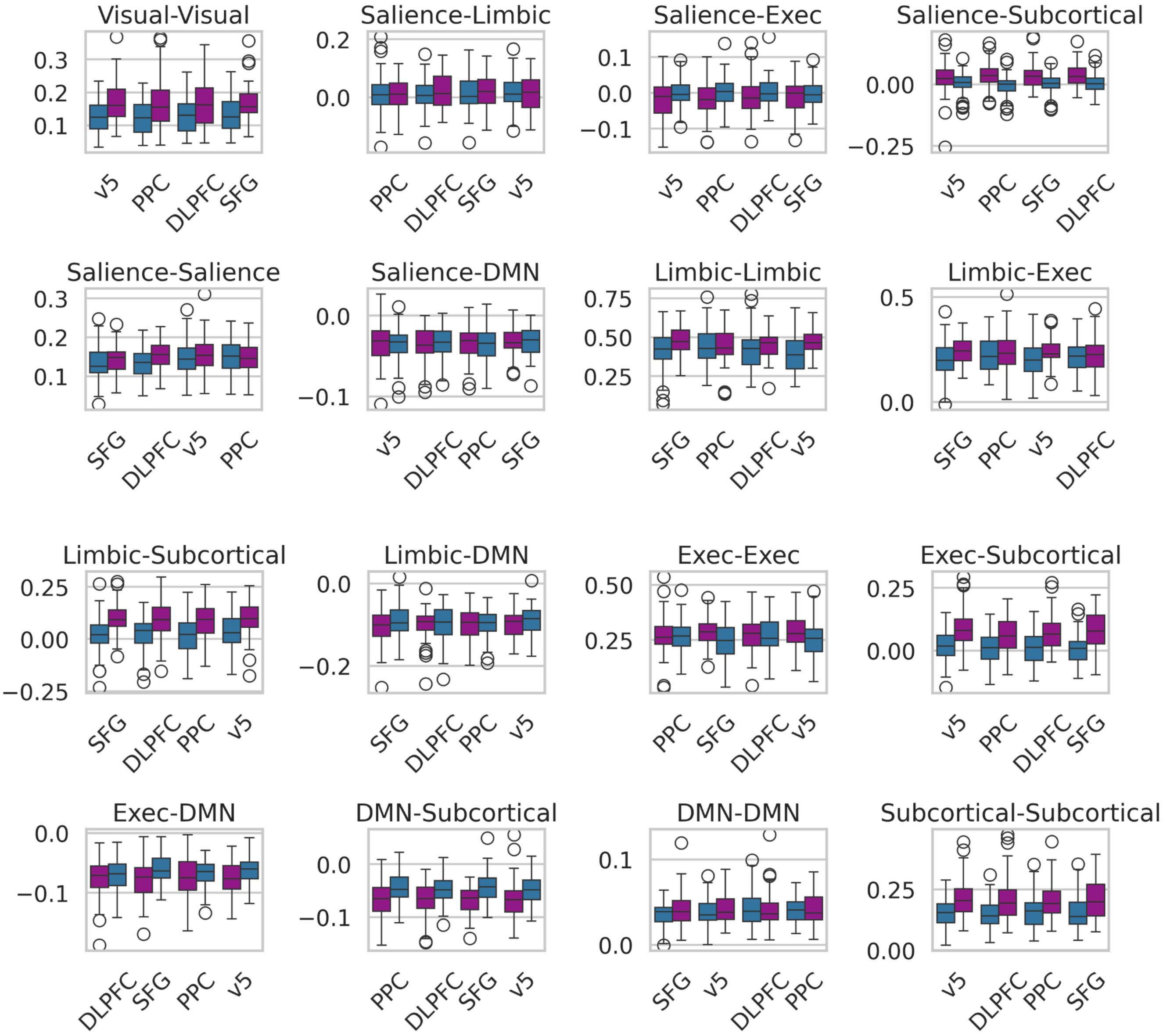
Visual-Visual
Salience-Limbic
Salience-Exec
Salience-Subcortical
Salience-Salience
Salience-DMN
Limbic-Limbic
Limbic-Exec
Limbic-Subcortical
Limbic-DMN
Exec-Exec
Exec-Subcortical
Exec-DMN
DMN-Subcortical
DMN-DMN
Subcortical-Subcortical
v5
PPC
DLPFC
SFG
